# Fixed dosing of tocilizumab in ICU admitted COVID-19 patients is a superior choice compared to bodyweight based dosing; an observational population pharmacokinetic and pharmacodynamic study

**DOI:** 10.1101/2021.05.10.21256933

**Authors:** Dirk Jan A.R. Moes, David J. van Westerloo, Sandra M. Arend, Jesse J. Swen, Annick de Vries, Henk-Jan Guchelaar, Simone A. Joosten, Mark G.J. de Boer, Teun van Gelder, Judith van Paassen

## Abstract

**Background:** Tocilizumab improves outcome, including survival, in intensive care unit (ICU) admitted COVID-19 patients. The currently applied dosage of 8 mg/kg is based on use of this drug for other indications, however is has not formally been investigated for COVID-19. In this study pharmacokinetics and dynamics of tocilizumab were investigated in ICU admitted COVID-19 patients.

**Methods:** This was an open-label, single-center observational pharmacokinetic and -dynamic evaluation study. Enrolled patients, with polymerase chain reaction confirmed Covid-19 were admitted to the ICU for mechanical ventilation or high flow nasal canula oxygen support. All patients were 18 years of age or older and received tocilizumab within 24 hours after admission to the ICU and received 6 mg dexamethasone daily as concomitant therapy.

**Results:** 29 patients were enrolled between 15 December 2020 and 15 March 2021. A total of 139 tocilizumab plasma samples were obtained covering the pharmacokinetic curve of day 0 up to day 20 after tocilizumab initiation. A population pharmacokinetic model with parallel linear and non-linear clearance was developed and validated. Average AUC_0-inf 1_^st^_DOSE_ was 938 [±190] ug/mL*days. Tocilizumab half-life was estimated to be 4·15 [±0·24] days. All patients had tocilizumab exposure above 1 ug/ml for at least 15 days.

**Conclusion:** This study provides evidence to support a fixed dose of 600 mg tocilizumab in COVID-19 patients. Furthermore our findings suggest that alternative cost saving regimens with even lower doses are likely to be as effective as the current 8 mg/kg recommendation.

**Funding:** No external funding was received for this work

**Background:** In the randomized controlled trial REMAP-CAP, the IL-6 receptor antagonist tocilizumab was shown to improve outcome, including survival in ICU admitted COVID-19 patients. Because obesity is a risk factor for development of severe COVID-19, concerns have been raised about overtreatment as well as undertreatment through weight-based dosing of tocilizumab. Furthermore pharmacokinetic and pharmacodynamic parameters of medications are often found to be different in severely ill patients when compared to mild or moderately ill patients. However, the effects of different dosing schedules were only investigated to a very limited extent in non-randomized observational studies. Hence, evaluation of the PK/PD parameters of tocilizumab in severely ill patients – is warranted.

**Added value of this study:** This study provides valuable information about the population pharmacokinetics and dynamics of tocilizumab in dexamethasone cotreated ICU admitted COVID-19 patients. This research shows that there is no rationale for the 8 mg/kg dosing recommendation in ICU patients. Fixed dosing of 600 mg tocilizumab is a cost saving, logistically attractive and safe alternative without losing efficacy.

**Implications of all the evidence:** Due to the ongoing pandemic, shortages of tocilizumab and other IL-6 receptor antagonists may be anticipated. A fixed tocilizumab dose regimen has many practical and safety advantages, e.g. it will reduce dosing errors and avoid unnecessary wastage of medication. More importantly, according to the data presented in this study, relative underdosing of patients with low, or low-normal bodyweight compared to patients with high bodyweight will be avoided. Last but not least, in view of the large number of patients currently being treated with these agents, a significant cost saving can also be expected.

## Introduction

The SARS-Cov-2 pandemic has caused over 145,000,000 coronavirus cases and over 3,100,000 deaths worldwide.^1^ The inflammatory storm associated with severe acute respiratory syndrome coronavirus 2 (SARS-CoV-2) infection damages the respiratory tract and causes high morbidity and mortality.^2^ Recently it was shown in the REMAP-CAP open label randomized controlled trial that next to dexamethasone, the treatment with interleukin-6 (IL-6) receptor antagonists tocilizumab and sarilumab significantly improved outcomes significantly, including survival in ICU admitted COVID-19 patients with acute respiratory distress syndrome (ARDS).^3-5^ Tocilizumab is the first marketed IL-6 blocking humanized antibody targeting IL-6 receptors and has proved its safety and effectiveness in therapy for rheumatoid arthritis (RA).^6, 7^ The current proposed dosage of 8 mg/kg tocilizumab (800 mg max) is based on the standard loading dose of RA, however evidence is lacking that this is also the optimal dose for COVID-19 ARDS.^8^ Tocilizumab is currently also registered for cytokine release syndrome (CRS) that can develop after CAR-T-cell therapy. Interestingly, the Food and Drug Administration (FDA) registration report consists of limited data of the CRS population and was further based on modelling and simulation studies based on data from RA patients. The FDA review committee concluded that as the maximum tolerated dose was not reached in previous trials for tocilizumab, the “safety threshold” from previous clinical experience was used to determine the acceptable dosage regimen and that the optimal dose and schedule are not well-established.^9^ Underdosing of tocilizumab might lead to suboptimal suppression of the hyperinflammation and overdosing might result in increased risk for secondary anti-microbial infections and unnecessary healthcare expenses. For optimization of the dosage regimen of tocilizumab, population pharmacokinetic and pharmacodynamic studies are essential. The pharmacokinetics and dynamics of tocilizumab in severe COVID-19 patients is still not well understood and no published data of tocilizumab pharmacokinetics and dynamics in this population is available. The pharmacokinetics of tocilizumab might be different in COVID-19 patients tocilizumab compared to RA and CRS patients. In addition, a wide range of inter-individual variability in the estimated pharmacokinetic parameters of tocilizumab has been demonstrated in RA, and this variability might even be higher in ICU admitted COVID-19 patients.^8^ The primary objective of this study was to characterize the population pharmacokinetics and pharmacodynamics of tocilizumab in ICU admitted COVID-19 patients. The secondary objective of this study was to identify potential covariates that influence the pharmacokinetics of tocilizumab and to develop an alternative dosing recommendation for tocilizumab ICU admitted COVID-19 patients.

## Methods

### Study design and participants

This study was an open-label, observational pharmacokinetic and pharmacodynamic evaluation study. Critically ill patients, 18 years of age or older, with PCR confirmed Covid-19 who were admitted to the ICU and receiving respiratory organ support were classified as having a severe disease state and were eligible for tocilizumab administration according to the local treatment protocol. Respiratory organ support was defined as either high flow oxygen support (flow rate > 40 liters, oxygen fraction > 0·4), or invasive or noninvasive mechanical ventilation. Tocilizumab was administered within 24 hours after starting organ support in the ICU. All patients were admitted at the Leiden University Medical Center, Leiden, The Netherlands between December 15^th^ 2020 and March 15^th^ 2021. Recruitment started on December 15^th^ 2020 and ended at February 15^th^ 2021. All patients had a follow up until 25 days after tocilizumab administration unless they were discharged or had died during the follow up period. Data and sample collection was performed between December 15^th^ 2020 and April 1th 2021. EDTA plasma samples were obtained from day 0 up to day 20 after tocilizumab administration. Furthermore demographic factors including gender, age, bodyweight, height, body mass index (BMI), body surface area (BSA) and routinely collected clinical chemistry laboratory values (serum creatinine, estimated glomerular filtration ratio CKD epi (eGFR), urea, total bilirubin, albumin, cytokine reactive protein (CRP), aspartate aminotransferase (ASAT), alanine aminotransferase (ALAT), gamma-glutamyl transferase (GGT), Ferritin, D-dimer) were obtained from the electronic healthcare record. The study was conducted in compliance with the Declaration of Helsinki and approved by the COVID-19 scientific and ethics committee of the Leiden University Medical Center with number coco 2020-033. Furthermore a waiver for the requirement for informed consent was granted.

### Procedures

For the evaluation of the pharmacokinetics of tocilizumab all time points from day 0 up to 20 days after dose administration were eligible for collection. Daily sampled leftovers from samples of the hematology tubes (K2-EDTA whole blood) were used to obtain plasma for free tocilizumab concentration determination. Plasma was obtained after centrifugation and stored at –20 °C until analysis. Tocilizumab concentrations were determined in the Sanquin Biologics Lab, Amsterdam, the Netherlands. Tocilizumab concentrations were determined with a validated ELISA-method using rabbit anti-tocilizumab antibodies to capture tocilizumab, and rabbit anti-tocilizumab F(ab’)2 fragments for detection as described earlier.^10, 11^ The lower limit of quantification (LLOQ) in plasma was 0·2 ug/mL; the overall precision and accuracy were 8% and 93%, respectively. Samples exceeding the upper limit of quantification of 250 ug/ml were diluted. Serum sIL-6R was determined by ELISA in duplicate using the commercial Quantikine Human IL-6sR kit (Bio-Techne Ltd. Abington, UK) as described earlier.^6^

### Outcomes

Primary endpoints were the pharmacokinetic and dynamic (CRP and sIL-6R) parameters of tocilizumab in ICU admitted COVID-19 patients. Next to the population pharmacokinetics parameters Clearance (CL), Distribution Volume (Vd), Maximum Elimination rate (Vmax), concentration at which the elimination pathway is half saturated (Km), AUC_0-inf 1_^st^_DOSE_, half-life and average concentration (C_avg_) were also calculated. Secondary endpoints were identification of potential covariates and development of alternative dosing schedules. An exploratory endpoint was the difference in these pharmacokinetic parameters in subgroups such as survivors and non-survivors. For the development of alternative dosing schedules Monte Carlo simulations in NONMEM using the final model were performed to assess whether the bodyweight dosing or fixed dosing was more appropriate to reduce variability in tocilizumab exposure. The predicted concentration time curves were simulated 1000 times for patients receiving 8 mg/kg for a body weight of 60, 70, 80, 90 and 100 kg compared to a fixed dose of 600 mg for all body weights categories. Furthermore predicted concentration time curves for fixed dosages of 800, 600, 400 and 200 mg and more personalized regimens were simulated.

### Statistical analysis and model development

The sample size calculation was based the sample size of the registration study of the cytokine release syndrome indication for tocilizumab approved by the FDA for. A nonlinear mixed effects model was developed to characterize the pharmacokinetics parameters of tocilizumab in ICU admitted COVID-19 patients using the first order conditional estimation with interaction approach. One- and two compartmental models with and without non-linear pharmacokinetics were evaluated. During model development, candidate models were evaluated for their decrease in Objective Function Value (OFV) calculated as the -2 log likelihood. A decrease in OFV of ≥6.63 was considered significant (Chi-squared [χ^2^], 1 degree of freedom [df], p<0.01).

Biological plausibility of a potential covariate was a selection criteria for the statistical assessment of covariates. Baseline demographic covariates (gender, age, bodyweight, BMI, BSA, and height) and laboratory covariates (creatinine, eGFR, urea, albumin, LDH, ASAT, ALAT, GGT, total bilirubin, albumin, CRP) were included in the covariate descriptive analysis. The influence of time-varying covariates was assessed for CRP, creatinine, eGFR, urea, albumin, LDH, ASAT, ALAT, GGT, total bilirubin and albumin. Covariates were subsequently entered in the univariate statistical analysis (Chi-squared [χ^2^], 1 degree of freedom [df], p<0.05) and subsequently in the cumulative forward inclusion/backward elimination procedure. A covariate was only retained if it led to a significantly improved fit (Chi-squared [χ^2^], 1 degree of freedom [df], p<0.01). Graphical displays based on the agreement between the observed and predicted drug concentrations and the uniformity of the distribution of the residuals were also considered for covariate inclusion. The final model was evaluated by means of a prediction corrected visual predictive check (VPC) based on 500 Monte-Carlo simulations. In addition, the precision of the parameter estimates was further assessed by means of a non-parametric bootstrap with resampling the dataset (n=1000 times). The population pharmacokinetic modelling was carried out using NONMEM v.7.4.4 and Perl Speaks NONMEM (v.5.0.0).^12-14^ Pirana was used for run interpretation (v. 2.9.8).^15^ R statistics (v. 3.4.4) was used for exploratory graphical analysis and for evaluation of the GOF and VPC.^16^

## Results

### Patients and samples

Twenty nine ICU admitted COVID-19 patients treated with 6 mg dexamethasone daily as well as 8 mg/kg tocilizumab were studied up to 25 days after tocilizumab administration. Patients had a mean age of 64 years (range 45-80 years). The majority of the patients were male (72·4 %). Mean total body weight was 96·4 kg (range 58-130 kg). All patients received concomitant anti-inflammatory treatment with dexamethasone 6 mg once daily for a maximum of 10 days during their hospital admission. 24 % received methylprednisolone in addition as rescue therapy when the initial course of dexamethasone and tocilizumab did not lead to clinical recovery after 10 days. Median time from Covid-19 first symptoms until tocilizumab administration was 11·7 days (range 1·6 - 24·6). All patients received 8 mg/kg tocilizumab with a maximum of 800 mg. Only 1 patient by accident received double dose. One other received the total dose of 8 mg/kg in 2 steps (within 12 hours) because the tocilizumab was temporarily out of stock. Detailed baseline characteristics of the studied population can be found in table 1. A total of 139 tocilizumab concentration-time points were obtained ranging from day 1 until day 20 after dose administration covering the entire pharmacokinetic curve. The mean and median number of samples per patient was 4·8 and 5 samples respectively with a range of 1 – 11 samples. All obtained data were used for the population pharmacokinetic analysis. In 80 samples evenly divided over all patients sIL-6R measurements could be performed.

**Table 1:** Baseline and clinical characteristics.

| Baseline characteristics | Mean | Range |
| --- | --- | --- |
| Male (%) | 72.4 |  |
| Age (yrs) | 64 | 45 - 80 |
| Bodyweight (kg) | 96.4 | 58.0 - 130.0 |
| Height (cm) | 177 | 165 - 192 |
| BMI | 30.9 | 20.1 - 46.1 |
| BSA | 2.2 | 1.7 - 2.5 |
| Smoking | 4 |  |
| Ureum (mmol/L) | 10 | 3.8 - 19.5 |
| Creatinine (umol/L) | 87 | 48 - 167 |
| eGFR (mL/min/1.73 m<sup>2</sup>) | 78 | 37 - 100 |
| Albumin (g/L) | 32 | 23 - 37 |
| LDH (U/L) | 565 | 197 - 1826 |
| CRP (mg/L) | 146 | 13.4 - 309 |
| ASAT (U/L) | 59 | 14 - 314 |
| ALAT (U/L) | 59 | 10 - 209 |
| GGT (U/L) | 125 | 16 - 1194 |
| Billirubin (total) (umol/L) | 12 | 3 - 42 |
| Ferritin (ug/L) | 1704 | 407 - 4441 |
| D-dimer (ng/mL) | 8737 | 333 - 83500 |
| Temperature (°C) | 36.6 | 35.3 - 39.2 |
| CT scan long infiltration (%) | 53 | 15 - 85 |
| CT score severeness (max 25) | 17.2 | 8 - 24 |
| Tocilizumab Dose administered (mg) | 781 | 472 - 1552 |
| Tocilizumab initiation (days after 1st symptoms) | 11.7 | 1.6 - 24.6 |
| Tocilizumab initiation (days after LUMC ICU admission) | 1.7 | 0.1 - 11.7 |
| Follow up (days) | 16.1 | 0.9 - 25.5 |
| Mechanical ventilation (%) | 82.8 |  |
| Ventilation in abdominal position (%) | 62.1 |  |
| SOFA score | 7.5 | 3 - 12 |
| APACHE IV score | 71.6 | 41 - 123 |
| SAPS III score | 43.9 | 37 - 52 |
| **Comorbidities (%)** |  |  |
| Diabetes | 6.9 |  |
| Hypertension | 41.4 |  |
| Cardiovascular disease | 31.0 |  |
| Chronic renal insufficiency | 6.9 |  |
| Cancer | 13.8 |  |
| COPD | 13.8 |  |
| **Anti-inflammatory comedication** |  | - |
| dexamethasone | 100 | - |
| methylprednisolone | 24.1 | - |
| hydrocortisone | 3.4 | - |

Tocilizumab plasma concentration data were best described by a one-compartment disposition model with parallel first order (linear) and Michaelis–Menten (nonlinear) elimination kinetics (figure 1). The diagnostic plots for the basic and final population pharmacokinetic model indicated that the observed and predicted data were in good agreement. Average CL was estimated to be 0·725 L/Day, average distribution of volume was 4·34 L. Vmax was 4·19 ug/day and Km was 0·22 ug/ml). Interindividual variability was identified for CL (18.9%) and Vd (21%). Residual variability was characterized by a combined error model with an additive error of 0·139 μg/ml and a proportional error of 17·1% (expressed as the coefficient of variation). An complete overview of the pharmacokinetic parameter estimates from the final population pharmacokinetic model in table 2. Average AUC_0-inf 1_^st^_DOSE_ was 938 [±190] ug/mL*days. Tocilizumab half-life was estimated to be 4·15 [±0·24] days. All patients had at least 15 days of tocilizumab exposure above 1 ug/ml(associated with complete IL-6 receptor blockade in RA and Castleman’s Disease).^6^ On average patients with lower bodyweight had lower exposure than patients with higher bodyweight as can be seen in figure 2.

**Figure 1.**
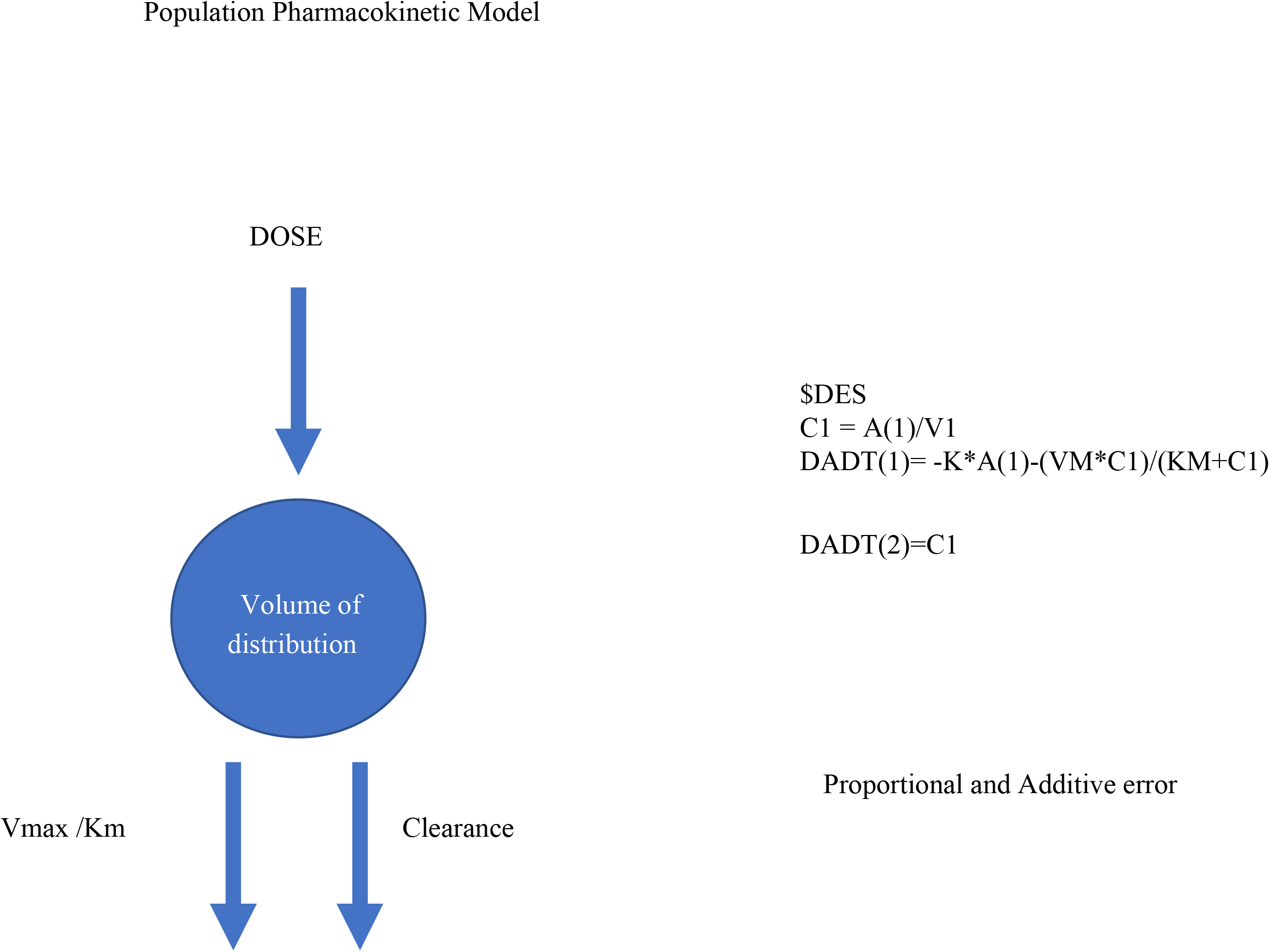
Schematic representation of the population pharmacokinetic model structure with parallel linear and non-linear clearance including differential equations.

**Table 2.**
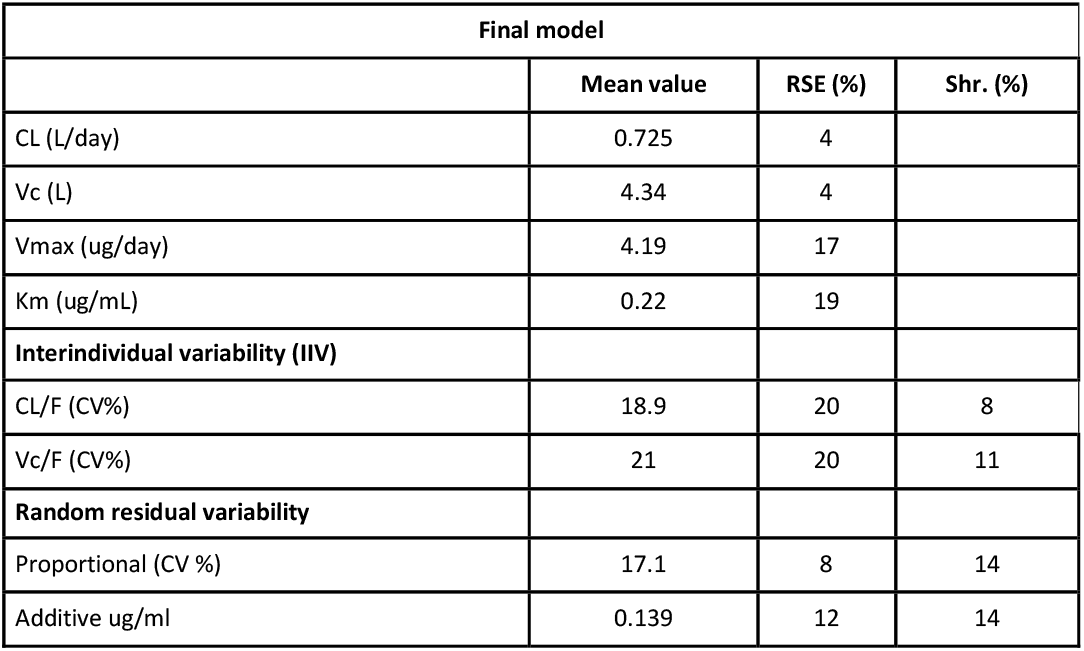
Population Pharmacokinetic parameters of the final model.

**Table 3A.** Pharmacokinetic and dynamic parameters differences between CRP relapse and non-relapse group.

| Parameter | NON CRP RELAPSE GROUP (N=22) |  | CRP RELAPSE GROUP (N=7) |  | TOTAL (N=29) |  |
| --- | --- | --- | --- | --- | --- | --- |
|  | Mean | (stdev) | Mean | (stdev) | Mean | (stdev) |
| AUC<sub>0-inf</sub> 1<sup>st</sup> DOSE (ug/mL*days) | 967 | 180 | 847 | 209 | 938 | 190 |
| Average concentration ug/ml *days | 45.9 | 7.8 | 39.6 | 9.5 | 44.0 | 9.5 |
| Half Life (days) | 4.20 | 0.20 | 4.01 | 0.33 | 4.15 | 0.24 |
| CRP baseline (mg/L) | 120.9 | 72.7 | 155.9 | 48.9 | 128.0 | 67.8 |

**Table 3B.** Pharmacokinetic and dynamic parameters differences between non-survivors and survivors.

| Parameter | Non-survivors (N=11) |  | Survivors (N=18) |  | Total (N=29) |  |
| --- | --- | --- | --- | --- | --- | --- |
|  | Mean | (stdev) | Mean | (stdev) | Mean | (stdev) |
| AUC<sub>0-inf</sub> 1<sup>st</sup> DOSE (ug/mL*days) | 928 | 238 | 944 | 162 | 938 | 190 |
| Average concentration ug/ml *days | 43.4 | 10.6 | 44.3 | 7.5 | 44.0 | 8.6 |
| Half Life (days) | 4.12 | 0.24 | 4.17 | 0.24 | 4.15 | 0.24 |
| CRP baseline (mg/L) | 121.9 | 56.8 | 131.0 | 75.2 | 128.0 | 67.8 |

**Figure 2.**
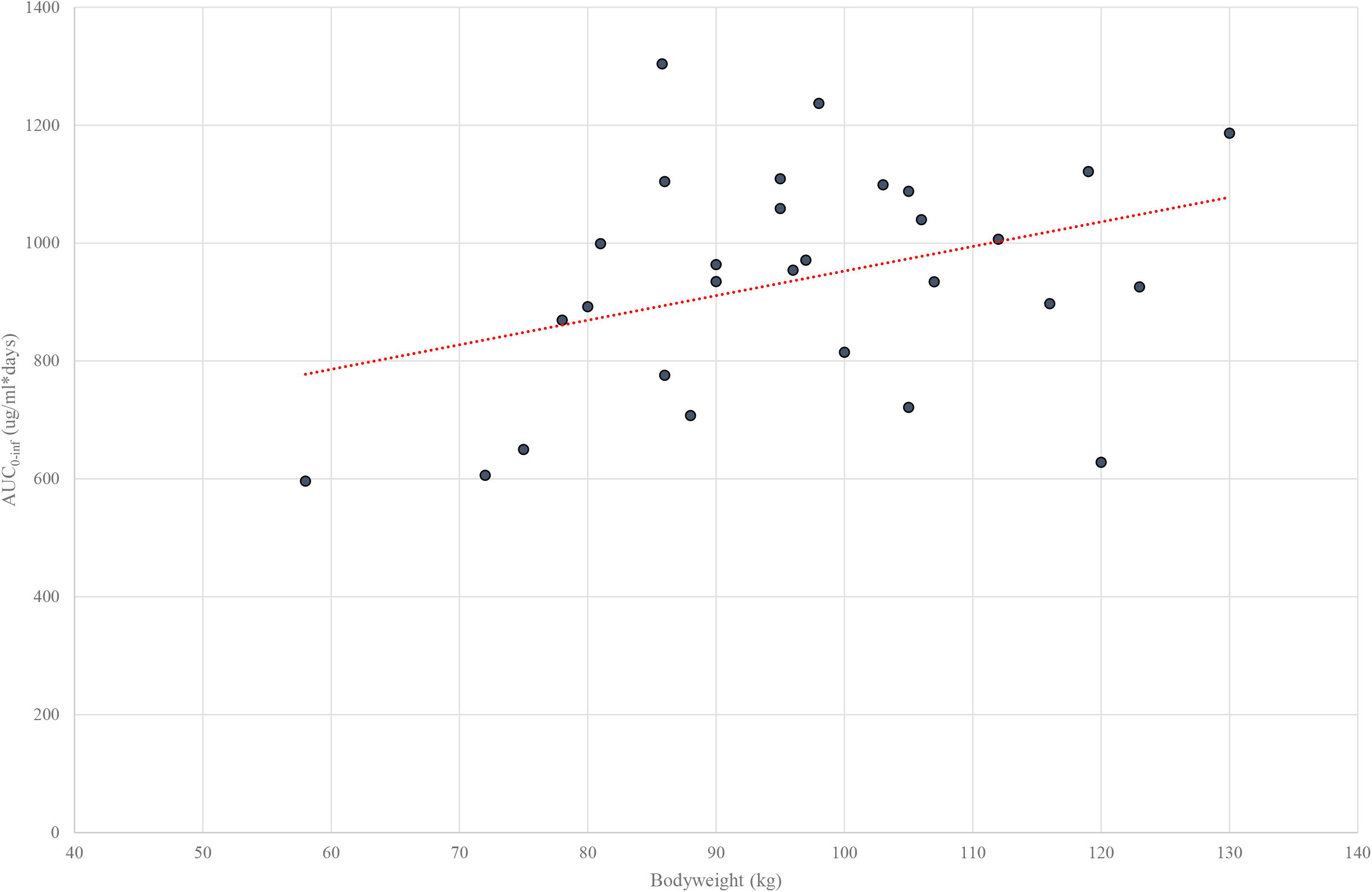
Graphical representation of individual tocilizumab exposure (AUC_0-inf 1_^st^_DOSE_) correlation with patients bodyweight with the current 8 mg/kg dosing recommendation.

The baseline demographic covariates (gender, bodyweight, age, BMI, BSA, height) and laboratory covariates (creatinine, eGFR, urea, albumin, LDH, ASAT, ALAT, GGT, total bilirubin, albumin, CRP) did not show any relationship between clearance or distribution volume after visual and descriptive analysis. After the stepwise covariate statistical analysis also no statistically significant covariates were identified. An extensive evaluation and model validation report including goodness of fit and the most important covariate evaluation plots can be found in supplementary file 1.

Figure 3A displays the individual predicted concentration-time curves, the measured tocilizumab concentration, the CRP value dynamics and the threshold of 1 ug/ml of all included patients. Figure 3B shows the individual predicted concentration-time curves, the measured tocilizumab concentration, the sIL-6R value dynamics and the threshold of 1 ug/ml of all included patients. Most variability in exposure is present in the first days after tocilizumab administration. In all patients CRP declined rapidly after tocilizumab administration and remained low for at least ten days. In a minority of patients (seven out of 29) an new rise in CRP was observed ten days after dose administration. Four of those patients were diagnosed with a secondary infection as described in the electronic healthcare record. As shown in figure 3A tocilizumab is detectable in all patients for at least 15 days after dose administration and for the majority of patients even longer. sIL-6R concentrations increased after tocilizumab administration and slowly declined after 17 days after administration. The group of patients with a relapse of CRP was more thoroughly compared to the non-CRP relapse group with regard to tocilizumab exposure and baseline CRP. The results are presented in Table 2A. No statistically significant differences in pharmacokinetic parameters were identified. Only a trend to higher baseline CRP was present in the CRP relapse group. A total of 11 of patients died during or after ICU admission. The majority (64%) of the patients who died did so within the first 25 days after tocilizumab administration. No statistically significant differences in tocilizumab pharmacokinetic parameters were found between survivors and non-survivors as presented in table 2B).

**Figure 3.**
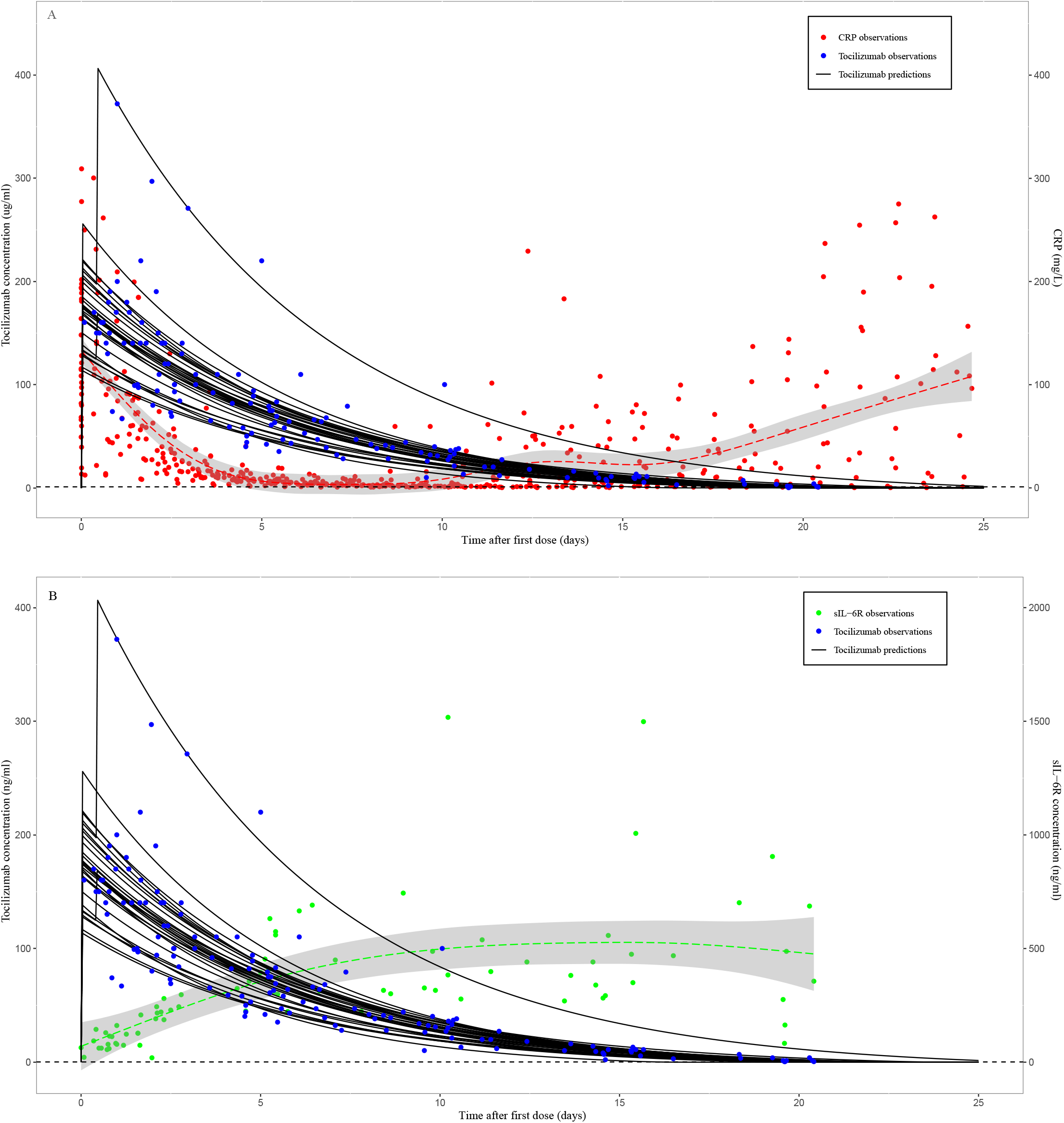
Top: Graphical representation of tocilizumab concentration time profiles (individual predictions), measured concentrations and CRP dynamics. Bottom: Graphical representation of tocilizumab concentration time profiles (individual predictions), measured concentrations and sIL-6r dynamics of all patients. The 1 ug/ml threshold of complete receptor saturation established in RA patients is presented with a dashed line.

Due to target mediated drug disposition the non-linear part of the clearance of tocilizumab is dominant in low concentration ranges and the linear part is dominant in the high concentration ranges.^8^ Based on the final model parameters the threshold for complete receptor saturation in this patient population was estimated to be 5 ug/ml as shown in supplementary file 1 (supplementary figure 4). To estimate the bodyweight coefficient, bodyweight was tested as a covariate to the model. The effect was not significant and the coefficient was estimated to be as low as 0.002. The final model was used to simulate the distribution of concentration time profiles of tocilizumab in patients weighing 60 up to 100 kg. In figure 4 the variability in concentration vs time curves is presented of 1000 simulated patients weighing between 60 and 100 kg receiving a 8 mg/kg tocilizumab dose versus all patients receiving a 600 mg fixed dose. Also the 1 ug/ml and 5 ug/ml receptor saturation thresholds are shown. Figure 4 shows that bodyweight based dosing increases variability in exposure compared to fixed dosing. Moreover with fixed dosing tocilizumab is above the 1 and 5 ug/ml threshold for a similar durations as bodyweight based dosing with a minimum of approximately 15 days. Figure 5 shows simulations in 1000 patients for the duration of tocilizumab exposure above these two thresholds, when using different alternative fixed dosages of 800, 600, 400, 200 mg. Even with a lowest fixed dose of 200 mg the tocilizumab exposure will be above these thresholds for at least 7 days in all patients. When dosing tocilizumab in severe COVID-19 patients one might want to realize a more personalized duration of tocilizumab exposure. Instead of giving a high dose once, multiple lower doses might be more suitable to personalize the tocilizumab treatment depending on the duration of the hyperinflammation period of the individual patient. Therefore different dosing strategies with the same cumulative dose were simulated and presented in figure 6. A single dose of 800 mg is compared to two doses of 400 mg with an interval of 10 days and four doses of 200 mg with an interval of 7 days. Repeated dosing with lower dosages (and lower cumulative dosages) results in longer durations of tocilizumab exposure above the saturation thresholds compared to a single high dose of 800 mg.

**Figure 4.**
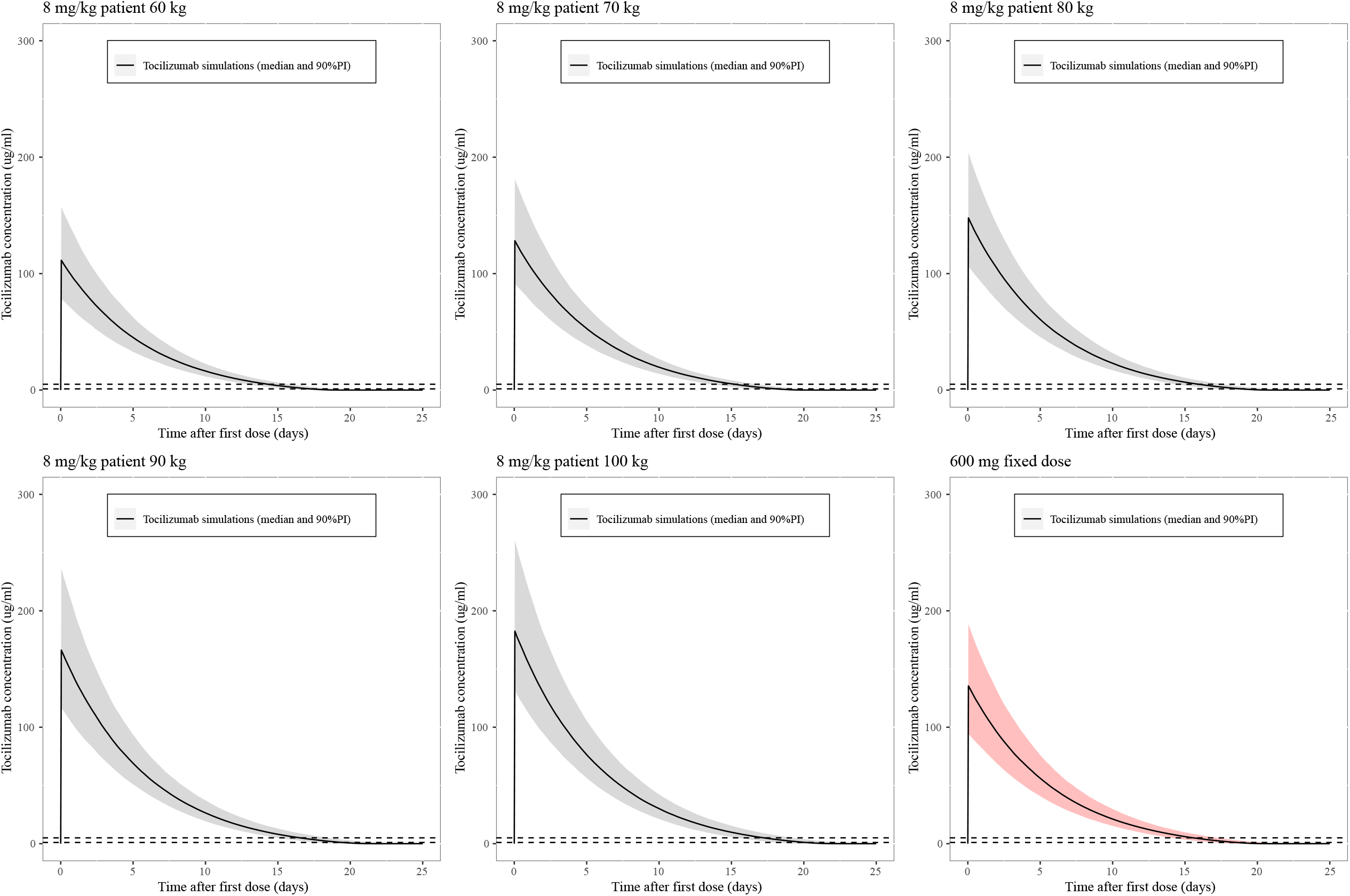
Variability in tocilizumab exposure when simulating 1000 patients of 60, 70, 80, 90 and 100 kg bodyweight receiving 8 mg/kg dosing versus 600 mg fixed dosing of tocilizumab. Median is presented with a solid black line and 90% prediction interval with a gray or red shaded area. The 1 and 5 ug/ml thresholds of complete receptor saturation established are presented with a black dashed line.

**Figure 5.**
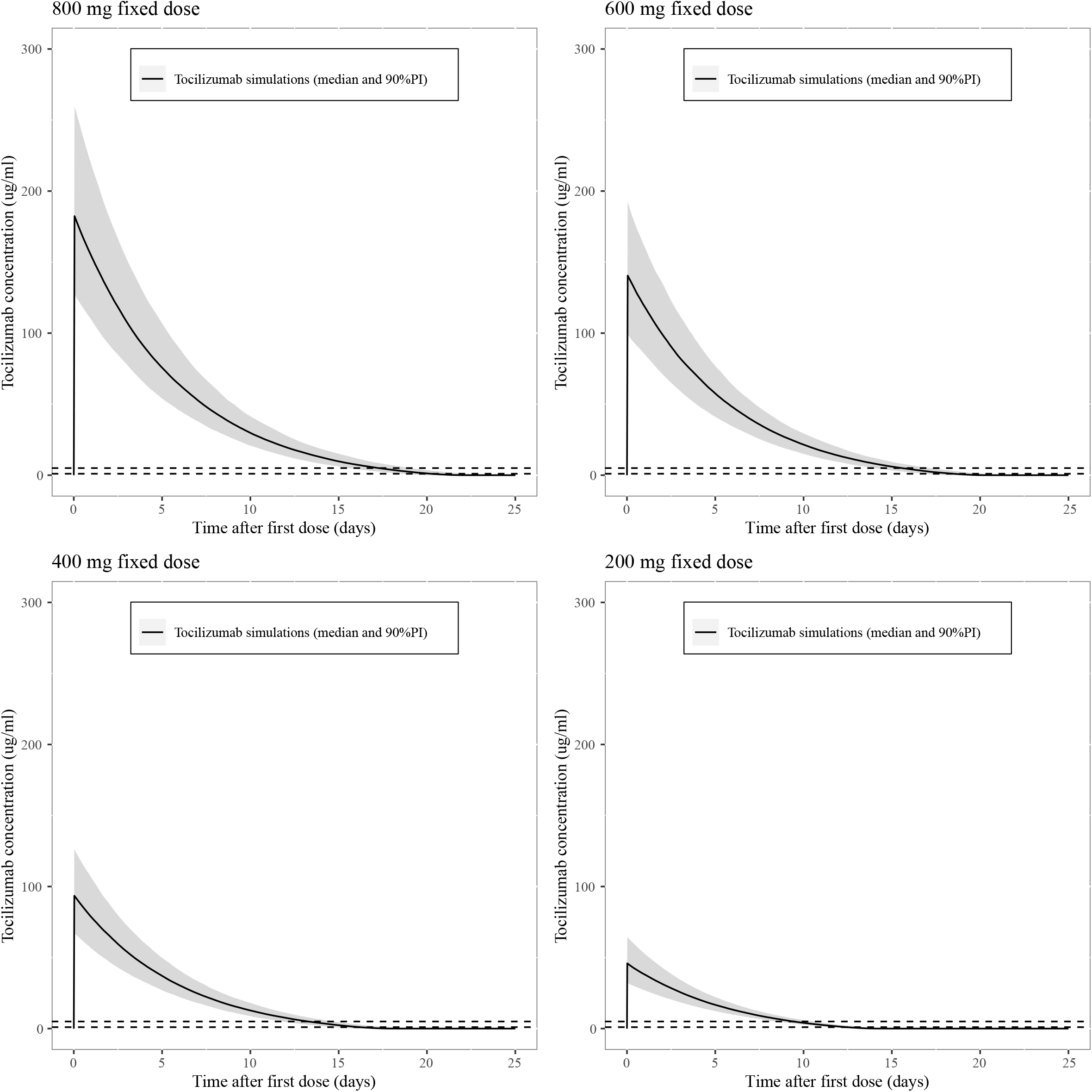
Variability in exposure when simulating 1000 patients receiving 800, 600, 400 or 200 mg fixed dose of tocilizumab. Median is presented with a solid black line and 90% prediction interval with a gray shaded area. The 1 and 5 ug/ml thresholds of complete receptor saturation established are presented with a black dashed line.

**Figure 6.**
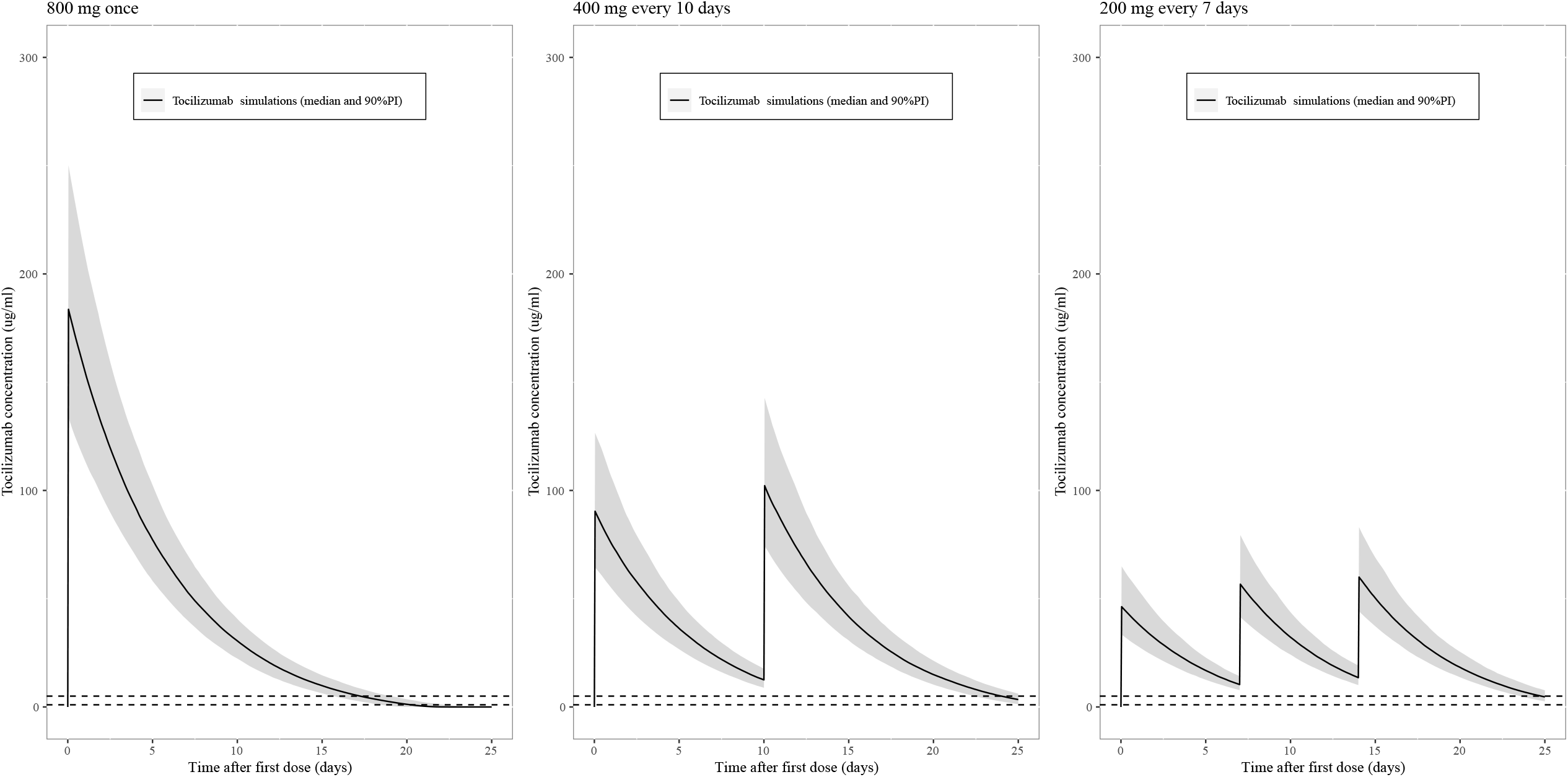
Simulated exposure of 1000 patients with alternative tocilizumab dosing strategies, 800 mg once, versus 400 mg every 10 days and 200 mg every 7 days. Median is presented with a solid black line and 90% prediction interval with a gray shaded area. The 1 and 5 ug/ml thresholds of complete receptor saturation established are presented with a black dashed line.

## Discussion

This is the first population pharmacokinetic and -dynamic study of tocilizumab in severe COVID-19 patients admitted to the ICU. The local treatment protocol, that included dexamethasone co-treatment for a maximum of 10 days, was based on the protocol used in the recently published REMAP-CAP trial.^4^ Our study shows that tocilizumab pharmacokinetics was variable in the investigated population and not dependent on body weight or other demographic and laboratory covariates. Previous studies have shown that patients with overweight are at risk for more severe course of COVID-19. Also in our population patients with overweight were overrepresented. Dosing tocilizumab based on bodyweight will lead to unnecessarily high tocilizumab concentrations in these patients. Most importantly our findings indicate that fixed dosing of tocilizumab will decrease interindividual variability in exposure compared to the current body weight based dosing while maintaining efficacy.

The tocilizumab plasma concentration–time courses were best described by an one-compartment pharmacokinetic model with parallel linear and nonlinear elimination, which is characteristic for monoclonal antibodies.^17^ A two compartment model which was identified earlier in RA with parallel first order (linear) and Michaelis–Menten (nonlinear) elimination kinetics was explored.^18^ However, this resulted in an unstable model with extremely high residual standard errors (%) of the pharmacokinetic parameters estimates. Although antibodies’ pharmacokinetics have usually been described using two-compartment models, when the sampling strategy is not intensive in the first day (the distribution phase), the peripheral compartment is not always identifiable.^19^ Our findings also show that tocilizumab clearance is faster than previously reported for rheumatic diseases (0·25 L/day) and in the CAR T-cell associated Cytokine Release Syndrome (0·5 L/Day).^9, 20^ This might be explained by different IL-6 mediated disease activity and different comedications used in COVID-19 patients compared to the other study populations.

The covariate analysis and simulation results confirm that a fixed-dosing approach for tocilizumab in severe COVID-19 patients reduces variability in exposure among weight categories compared to the current weight based dosing approach. Wang et al. claimed that the best dosing approach for monoclonal antibodies is dependent on the steepness of the relationship between CL and weight.^17^ In our study, this steepness was estimated to be 0.002, which is far below this 0.5 above which bodyweight dosing would be rational. This is in line with the findings of Bastida et al. and reconfirms our hypothesis that fixed dosing is a better dosing strategy to achieve a more consistent exposure among all weight groups^20^. The absence of clear relationships with the other demographic and laboratory covariates are in line with previous findings in RA patients when taking into account the specific population characteristics of current cohort.^8, 20^

In patients with RA and Castleman’s disease Nishimoto et al. showed that as long as free tocilizumab was detectable, meaning at a concentration of 1 ug/ml, sIL-6R was saturated with tocilizumab and that IL-6 signaling was completely inhibited.^6^ Our own analysis suggest a threshold of 5 ug/ml which is close to the 1 ug/ml threshold. Whether these findings can be extrapolated to all used treatment protocols of COVID-19 patients warrants further investigation. This would indicate that even doses below 600 mg are as effective as the current 8 mg/kg dose. This hypothesis is supported by findings that a fixed dose of 400 mg intravenous tocilizumab also leads to rapid CRP decline and improved survival compared to controls that were not treated.^21^ Furthermore a retrospective cohort study showed that subcutaneous administration of 324 mg tocilizumab showed comparable results to 8 mg/kg intravenous administration.^22^ The timing of tocilizumab administration seems to be the most important factor for success as shown in the REMAP-CAP trial and the dose can still be optimized.^4^ In cases with shorter duration of the hyperinflammation period doses can be possibly omitted, with the advantage of reducing risk of superinfections and savings on drug expenses.

Since the RECOVERY and REMAP-CAP open label randomized trials have demonstrated an effect of tocilizumab on COVID-19 associated mortality. Treatment with IL-6 receptor antagonists has now been embedded in national and international treatment guidelines.^23, 24^ However, the effects of different dosing schedules have not been investigated in these trials. Because obesity is a risk factor for development of severe COVID-19, concerns have been raised about overtreatment as well as undertreatment through weight-based dosing. Due to the ongoing pandemic shortages of tocilizumab and other IL-6 receptor antagonists may be anticipated upon. A fixed dosed regimen has many practical advantages, including a reduction in dosing errors and avoidance unnecessary wastage of medication. Furthermore, according to the data presented in this study, relative underdosing of patients with low, or low-normal bodyweight will be avoided.

Our study has some limitations. Due to the observational study design and unpredictable course of COVID-19 patients during ICU admission sampling could be not performed in an equal manner for all patients. Nevertheless the population approach used in the analysis allows to draw solid conclusions from unevenly distributed datasets. Furthermore a total of no less than 139 samples resulted in reliable pharmacokinetic parameter estimates. The sample size is relatively low, but similar to the dataset that was required (30 patients 140 samples) to obtain a FDA registration for CRS of tocilizumab, and model performance was good.^9^

In conclusion, our findings strongly support fixed dosing of tocilizumab in ICU admitted COVID-19 patients with a dose of 600 mg, as it leads to less variability in exposure compared to the currently applied bodyweight dosing. In patients with low bodyweight the fixed dose will avoid relative under-exposure, while in patients with high bodyweight the fixed dosing strategy will avoid unnecessary overexposure and excessive costs. Our findings also suggest that alternative cost saving regimens with even lower doses than 600 mg are likely to be as effective.

## Data Availability

Manuscript is currently under review at a journal. Anonimized data is available upon reasonable request.

## Acknowledgements

We are indebted to all the patients with COVID-19 who participated in this research. We sincerely thank all medical students, laboratory staff and clinical staff of the Leiden University Medical Center who, until this time, contributed to obtaining the data. We would like to thank Marjolein van Wolfswinkel (Department of Infectious Diseases) for performing the sIL-6R measurements.

