## Supplementary File 1 for "Fixed dosing of tocilizumab in ICU admitted COVID-19 patients is a superior choice compared to bodyweight based dosing; an observational population pharmacokinetic and pharmacodynamic study"

### Extensive model evaluation and validation

The statistical distributions of the population pharmacokinetic parameter estimates obtained from the bootstrap analysis are shown in supplementary table 1. The median values of the parameters estimated from the bootstrap analysis were in good agreement with the point estimates and the 95% CIs were reasonably narrow, demonstrating acceptable precision. Furthermore, the model parameters had adequately low levels of η-shrinkage for CL (8%) and V (11%). Visual and numerical predictive checks demonstrated good predictive performance of the final population pharmacokinetic model (Supplementary figure 1). Furthermore, the goodness of fit plots show good model performance (Supplementary figure 2)

**Supplementary table 1: Population pharmacokinetic parameters of final model and bootstrap results**

| **Final model** | | | | **1000 Bootstrap runs results** | |
|---|---|---|---|---|---|
| | Mean value | RSE (%) | Shr. (%) | Median Value | 95% CI |
| CL (L/day) | 0.725 | 4 | | 0.72 | 0.64 - 0.80 |
| Vc (L) | 4.34 | 4 | | 4.37 | 3.97 - 4.78 |
| Vmax (ug/day) | 4.19 | 17 | | 4.49 | 2.88 - 8.24 |
| Km (ug/mL) | 0.22 | 19 | | 0.30 | 0.02 - 3.17 |
| **Interindividual variability (IIV)** | | | | | |
| CL/F (CV%) | 18.9 | 20 | 8 | 18.2 | 9.5 - 26.3 |
| Vc/F (CV%) | 21 | 20 | 11 | 19.4 | 10.0 - 28.3 |
| **Random residual variability** | | | | | |
| Proportional (CV %) | 17.1 | 8 | 14 | 17.2 | 13.1 - 20.3 |
| Additive ug/ml | 0.139 | 12 | 14 | 0.1 | 0.0 - 0.8 |

**Supplementary Figure 1: Prediction Corrected Visual Predictive Check**

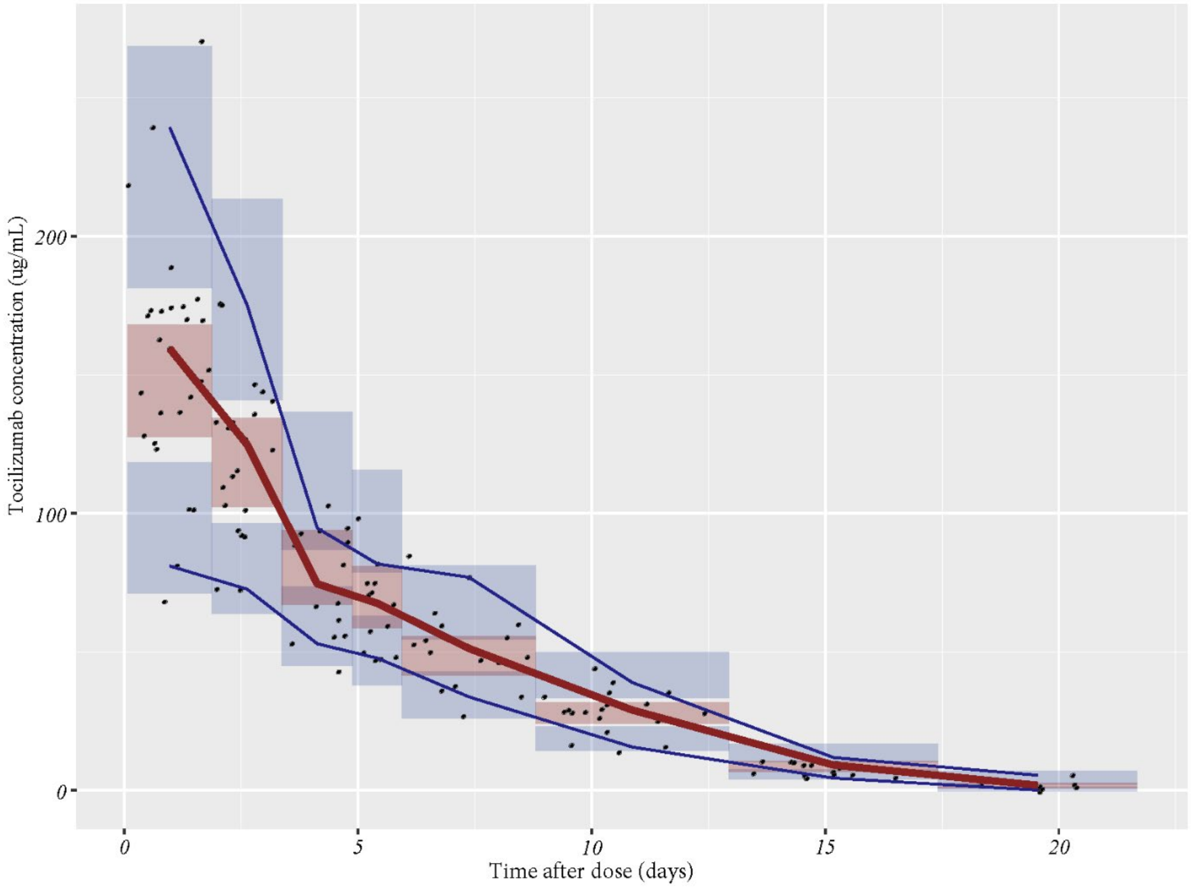


Prediction corrected visual predictive check of the final pharmacokinetic model. Comparisons were performed between the 10th. 90th (blue solid) and 50th (red solid line) percentiles of the observed tocilizumab plasma concentrations (closed circles) vs. time after first dose (days) and the 80% confidence interval (shaded area) obtained from 500 simulations.

**Covariates plots**

As shown in supplementary figure 3 no correlation between tocilizumab clearance and bodyweight, BSA, BMI, CRP at baseline, Height or Age could be identified. The loess line is almost flat over the whole range for every covariate. This was also statistically confirmed.

**Ratio between linear clearance, non-linear clearance (target mediated drug disposition) and total clearance of the total tocilizumab concentration range.**

Supplementary figure 4 shows that target mediated clearance (non-linear clearance) represented only a small portion of the total clearance over the majority of the tocilizumab concentration range that was observed in ICU admitted COVID-19 patient population. At concentrations above 5 ug/ml the linear clearance (non-target mediated clearance) is dominant. This means that certainly above a tocilizumab concentration of 5 ug/ml the target mediated pathway is completely saturated.

Supplementary figure 2: Goodness of fit plots

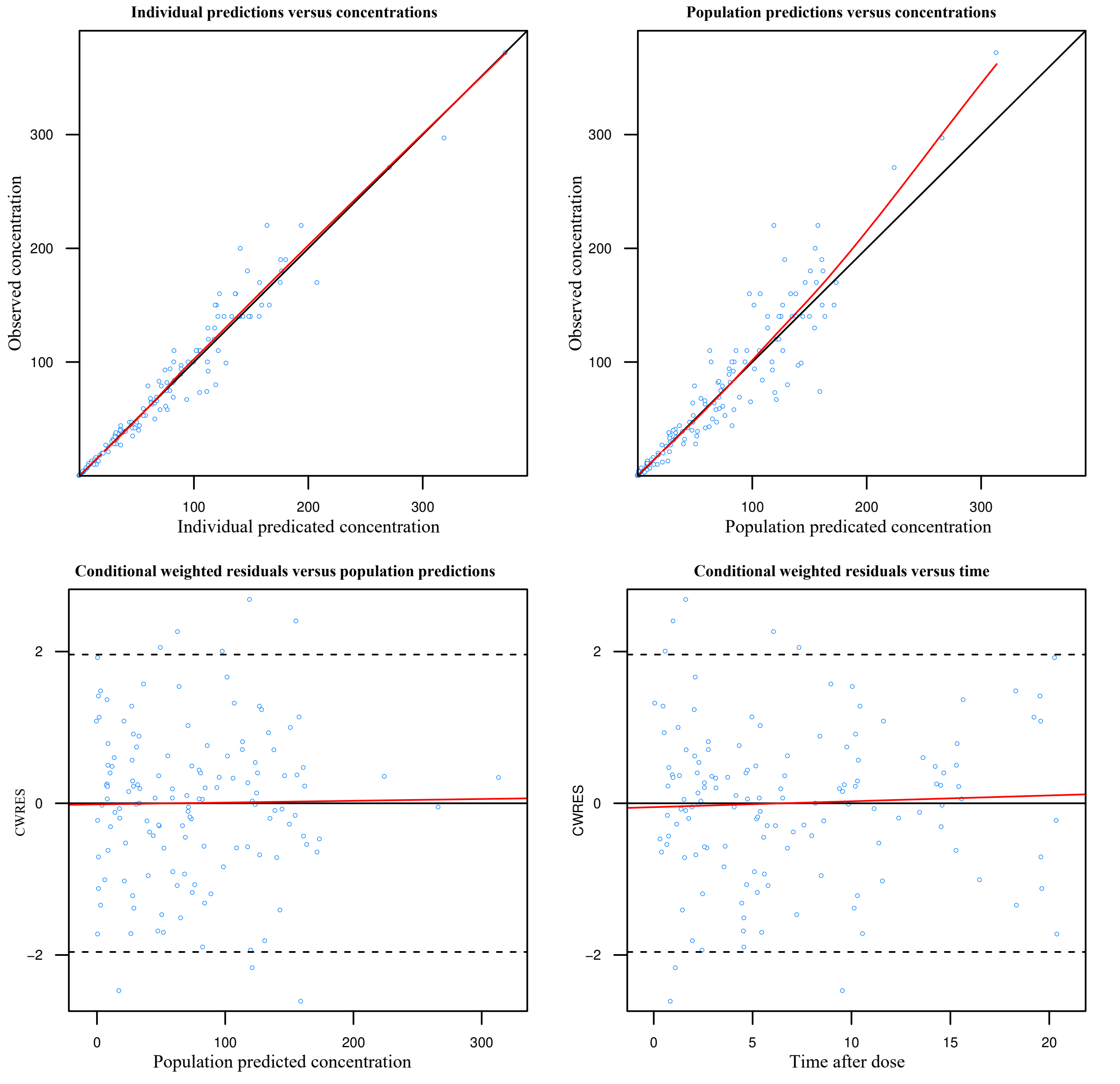

Supplementary figure 3: Covariate Plots

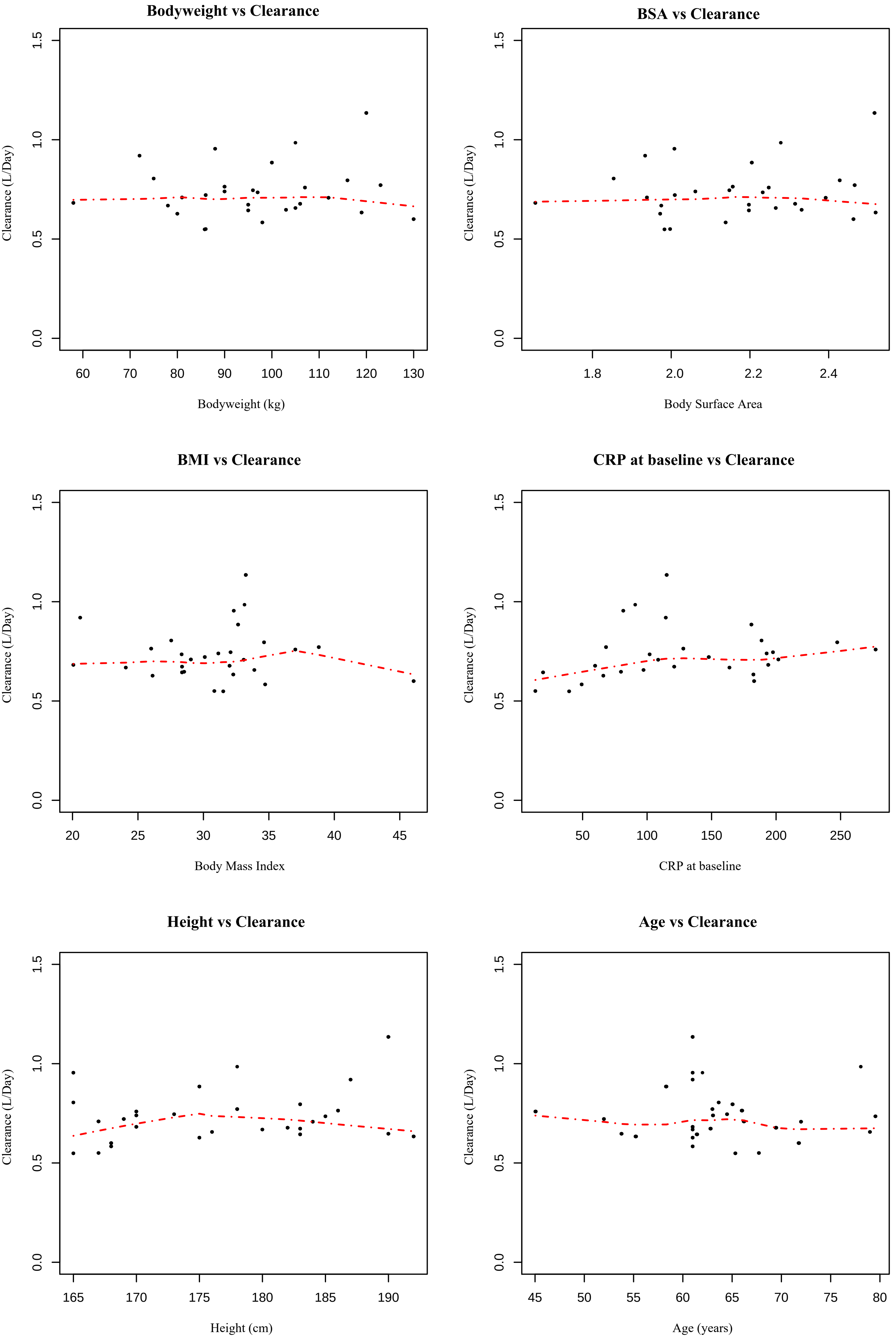

Supplementary figure 4: Ratio linear, non-linear and total clearance (CL) over the concentration range

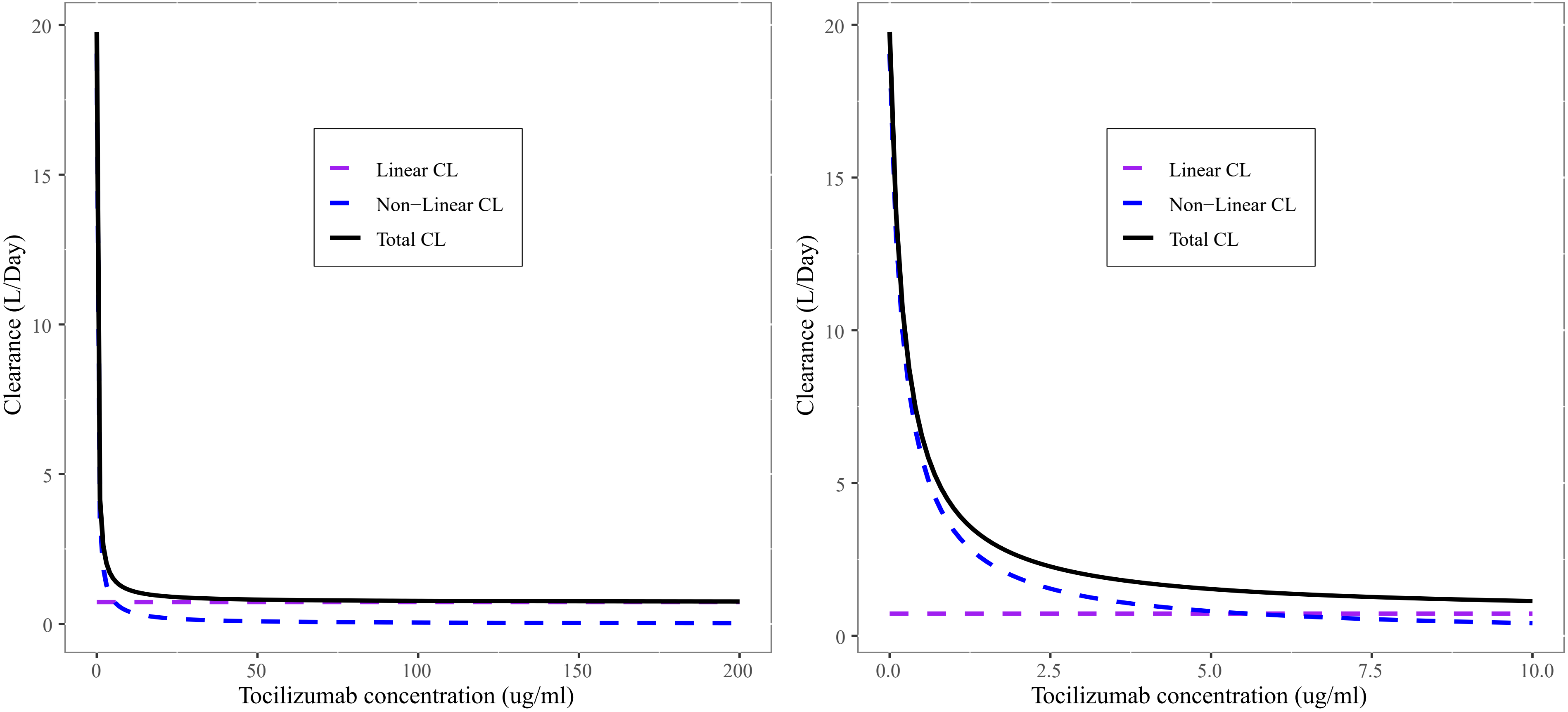
